# Associations of the Triglyceride and Glucose Index with Hypertension Stages, Phenotypes, and Progressions among Middle-aged and Older Chinese

**DOI:** 10.1101/2022.08.18.22278942

**Authors:** Shiyi Shan, Shuting Li, Keyao Lu, Jin Cao, Weidi Sun, Jiali Zhou, Ziyang Ren, Siyu Zhu, Leying Hou, Dingwan Chen, Peige Song

## Abstract

**Background:** The triglyceride and glucose (TyG) index has been proposed as a surrogate indicator of insulin resistance. By far, the associations of the TyG index with hypertension stages, phenotypes, and progressions remain unclear.

**Methods:** The data originated from two waves (2011 and 2015) of the China Health and Retirement Longitudinal Study (CHARLS). Participants with systolic blood pressure ≥ 140 mmHg and/or diastolic blood pressure ≥ 90 mmHg and/or using antihypertensive medications were considered hypertensive. After excluding those under antihypertensive medications, hypertension stages were classified as stage 1 and stage 2, and phenotypes were classified as isolated systolic hypertension (ISH), isolated diastolic hypertension (IDH), and systolic diastolic hypertension (SDH). Multinomial logistic regression was used to investigate the associations of the TyG index with hypertension stages and phenotypes, together with their progressions from 2011 to 2015.

**Results:** At baseline in CHARLS 2011, a total of 8,209 participants were recruited, among whom 3,169 (38.6%) were hypertension. Compared with individuals with the lowest quartile (Q1) of TyG index, those with the highest quartile (Q4) were significantly associated with increased risks of stage 1 hypertension (odds ratio [OR] 1.71, 95% confidence interval [CI] 1.38-2.13), stage 2 hypertension (1.74, 1.27-2.38), ISH (1.66, 1.31-2.11), IDH (2.52, 1.26-5.05), and SDH (1.65, 1.23-2.23). Similar results were found when the TyG index was used as a continuous variable. From 2011 to 2015, a higher baseline TyG index was revealed to be significantly associated with the progressions from normotension to stage 1 (for Q4 vs Q1: 1.45, 1.05-2.00; for per-unit: 1.39, 1.16-1.65), normotension to ISH (for per-unit: 1.28, 1.04-1.56), and normotension to IDH (for Q4 vs Q1: 3.46, 1.42-8.44; for per-unit: 1.94, 1.27-2.97).

**Conclusions:** The TyG index was significantly associated with different hypertension stages, phenotypes and their progressions. Our findings highlight the importance of the TyG index as a potential surrogate indicator for early hypertension screening and management.

## Introduction

Hypertension can lead to myocardial infarction, renal failure, and even death if not treated appropriately^1,2^. According to the World Health Organization report, hypertensive heart disease is one of the top 10 causes of death in upper-middle-income countries in 2019^3^. It is estimated that 1.28 billion adults aged 30-79 years are affected by hypertension in 2021^4^. In China, hypertension has evolved into a serious public health concern, as its burden grows in tandem with increased urbanization, population ageing, and exposure to unhealthy lifestyles^5-10^. A review from Yin et al. reported that the prevalence of hypertension ranged from 18.0% to 44.7% in China^11^. According to the *Chinese Guidelines for Prevention and Treatment of Hypertension (2018 Edition)*, hypertension statuses can be divided into hypertension and normotension, hypertension stages are classified as stage 1, stage 2, and stage 3 and hypertension phenotypes are classified as isolated systolic hypertension (ISH), isolated diastolic hypertension (IDH), and systolic diastolic hypertension (SDH)^12^. Besides, as the course of hypertension progresses, changes in hypertension statues or stages or phenotypes will occur, namely hypertension progressions. Investigating the progressions of hypertension can help to conduct dynamic management and treatment of hypertension more accurately.

The triglyceride and glucose (TyG) index, calculated as *ln*(*fasting triglycerides* [*TG*] [*mg/dl*] × *fasting plasma glucose*[*FPG*][*mg/dl*]/2), has attracted increasing interest due to its excellent sensitivity (79%) and specificity (86%) for diagnosing insulin resistance (IR)^13-15^. Previous studies mainly focused on the association between the TyG index and the risk of hypertension in adults, and suggested that the TyG index was a superior indicator for hypertension than independent lipid or glycemic markers^16-19^. To the best of our knowledge, limited studies have explored the associations of the TyG index with various hypertension stages and phenotypes, let alone their progressions.

To fill the research gap, this study aims to evaluate the associations of the TyG index with hypertension stages and phenotypes. And we also investigate whether the TyG index is associated with the progressions of hypertension stages and phenotypes.

## Methods

### Study population

This study used data from the China Health and Retirement Longitudinal Study (CHARLS), a nationwide longitudinal survey of people aged 45 years or above^20^. A stratified four-stage probability sampling strategy was adopted to recruit participants from 150 counties and urban districts across 28 provinces in China. The baseline survey was conducted in 2011. Respondents were then followed up with a face-to-face computer-assisted personal interview every two years (2013, 2015, and 2018, respectively). Detailed study procedures can be found elsewhere^20^. CHARLS was approved by the Behavioral and Social Research division of the National Institute on Aging of the National Institute of Health, the Natural Science Foundation of China, the World Bank, and Peking University.

### Measurement of blood-based biomarkers

Blood sample collection was done once every two follow-up periods (2011 and 2015). Biomarkers were measured using overnight fasting blood samples, which were promptly transported to the local laboratory and stored at 4□. The blood samples were centrifugated and stored at −20□ before being transported to the China Center for Disease Prevention and Control (CDC) in Beijing and frozen at −70□ before analysis^20^. Routine blood tests were performed at township hospitals or the local CDC. FPG was measured using the enzymatic method. TG, high-density lipoprotein cholesterol (HDL-C), and low-density lipoprotein cholesterol (LDL-C) were measured using the enzymatic colour metric method^21^. C-reactive protein (CRP) was measured using the colourimetric enzyme-linked immunosorbent assay^22^.

The TyG index was calculated as *ln*[*TG* (*mg/dl*) × *FPG* (*mg/dl*)/2)]^13^.

### Definition of hypertension stages, phenotypes and progressions

Blood pressure (BP) was measured three times in a sitting position at a 45-second interval using an electronic monitor (Omron model HEM-7200). The average of the three measurements was calculated to the nearest 0.1 mmHg for analysis. Hypertension statuses were divided into hypertension and normotension. Hypertension was defined as mean systolic blood pressure (SBP) ≥ 140 mmHg and/or mean diastolic blood pressure (DBP) ≥ 90 mmHg and/or using antihypertensive medications^12,20^. Normotension was defined as SBP < 140 mmHg and DBP < 90 mmHg. After excluding those with antihypertensive medications, hypertension stages were further classified as stage 1 hypertension (SBP ≥ 140 mmHg and < 159 mmHg and/or DBP ≥ 90 mmHg and < 99 mmHg), stage 2 hypertension (SBP ≥ 160 mmHg and < 179 mmHg and/or DBP ≥ 100 mmHg and < 109 mmHg), and stage 3 hypertension (SBP ≥ 180 mmHg and/or DBP ≥ 110 mmHg). Given the sample size constraints, we combined stage 2 and stage 3 hypertension into stage 2 hypertension. Hypertension phenotypes were also classified as ISH (SBP ≥ 140 mmHg and DBP < 90 mmHg), IDH (SBP < 140 mmHg and DBP ≥ 90 mmHg), and SDH (SBP ≥ 140 mmHg and DBP ≥ 90 mmHg). More details are shown in Table S1. The progressions of hypertension statuses were classified as maintained normotension, normotension to hypertension, and maintained hypertension. The progressions of hypertension stages were classified as maintained normotension, normotension to stage 1, normotension to stage 2, maintained stage 1, stage 1 to stage 2, and maintained stage 2. The progressions of hypertension phenotypes were classified as maintained normotension, normotension to ISH, normotension to IDH, normotension to SDH, maintained ISH, ISH to SDH, and maintained SDH.

### Definition of covariates

Anthropometric measurements, including height, weight, and waist circumstance (WC), were made at every 2-year follow-up. Information on age, sex, residence, education, economic status, tobacco use, and alcohol consumption were collected through questionnaires. The residence was classified as urban and rural according to the location of the participants. Education was divided into illiterate, primary school and below, and junior high school or above. Economic status was evaluated by the natural logarithm of per capita expenditures (ln[PCE])^23^. The bottom, middle, and top tertile of ln(PCE) indicated poor, middle, and rich economic status, respectively. Tobacco use was categorized as never smoking and ever smoking (including current smoking). Likewise, alcohol consumption was classified as never drinking and ever drinking (including current drinking). Body mass index (BMI) was calculated as (weight [kg] / [height [m]]^2^). The categories of general obesity included normal (BMI<24 kg/m^2^), overweight (BMI≥24 kg/m^2^ and BMI<28 kg/m^2^), and obesity (BMI≥28 kg/m^2^)^24^.

### Statistical analysis

Characteristics of participants at baseline (CHARLS 2011) were described in the form of the median with interquartile range (IQR) for continuous variables with skewed distribution, and number with percent (%) for categorical variables. The basic characteristics of participants between the included participants and excluded participants, between normotension and hypertension, among different hypertension stages, hypertension phenotypes, and hypertension progressions were compared using Wilcoxon rank-sum test for continuous variables and the Chi-square test for categorical variables.

The included participants were divided into four groups based on their TyG index levels (quartile 1, quartile 2, quartile 3 and quartile 4). In the data analysis, the TyG index was considered as a continuous (per-unit) variable and converted to a categorical (quartiles) variable.

For the cross-sectional analysis in CHARLS 2011, we used multinomial logistic regression to investigate the associations of the TyG index with hypertension statuses, stages, and phenotypes, with normotension as reference. The model was adjusted for age, sex, residence, education, economic status, tobacco use, alcohol consumption, general obesity, WC, LDL-C, HDL-C, and CRP. Meanwhile, considering the sex-TyG index interaction, we conducted sex-stratified multinomial logistic regression as well^25^.

For the longitudinal analysis using CHARLS 2011 and 2015, we excluded those who had taken antihypertensive medications to ensure proper classification of hypertension stages and phenotypes. Participants interviewed in both waves with complete data on SBP and DBP were included to further investigate the associations between the TyG index and hypertension progressions. Those who had non-attenuated hypertension progression patterns including maintained normotension, normotension to hypertension, and maintained hypertension based on their hypertension statuses from 2011 to 2015 were included. Subsequently, we selected participants who had non-attenuated hypertension stages (i.e. maintained normotension, normotension to stage 1, normotension to stage 2, maintained stage 1, stage 1 to stage 2, and maintained stage 2) and phenotypes (i.e. maintained normotension, normotension to ISH, normotension to IDH, normotension to SDH, maintained ISH, ISH to SDH, and maintained SDH) to explore whether the TyG index was associated with hypertension stages and phenotypes progressions, respectively. All of these analyses were conducted using sex-stratified multinomial logistic regression with maintained normotension as reference.

This study was done following the Strengthening the Reporting of Observational Studies in Epidemiology (STROBE) guidelines^26^. SAS statistical software (version 9.4; SAS Institute Inc., Cary, NC, USA) was used. All analyses were two-sided, and statistical significance was defined as a *P* value <0.05.

## Results

### Selection of study participants

A total of 17,708 participants were recruited at baseline in CHARLS 2011. After excluding those under the age of 45 (n=289) or with incomplete data on sex, education, economic status, tobacco use, SBP, DBP, weight, height, and WC (n=6,018), who had incomplete data on TG, FPG, HDL-C, and LDL-C (n=3,192), finally 8,209 participants were included in analyses (Figure 1). Characteristic comparisons between the included (n=8,209) and excluded (n=9,499) participants were shown in Table S2. Table S3 showed that of the 8,209 individuals included in CHARLS 2011, 5,040 (61.4%) had normotension, and 3,169 (38.6%) had hypertension. Significant differences in age, residence, education, economic status, alcohol consumption, general obesity, WC, LDL-C, HDL-C, CRP, and the TyG index between different hypertension statuses were observed (all *P* values <0.05).

**Figure 1.**
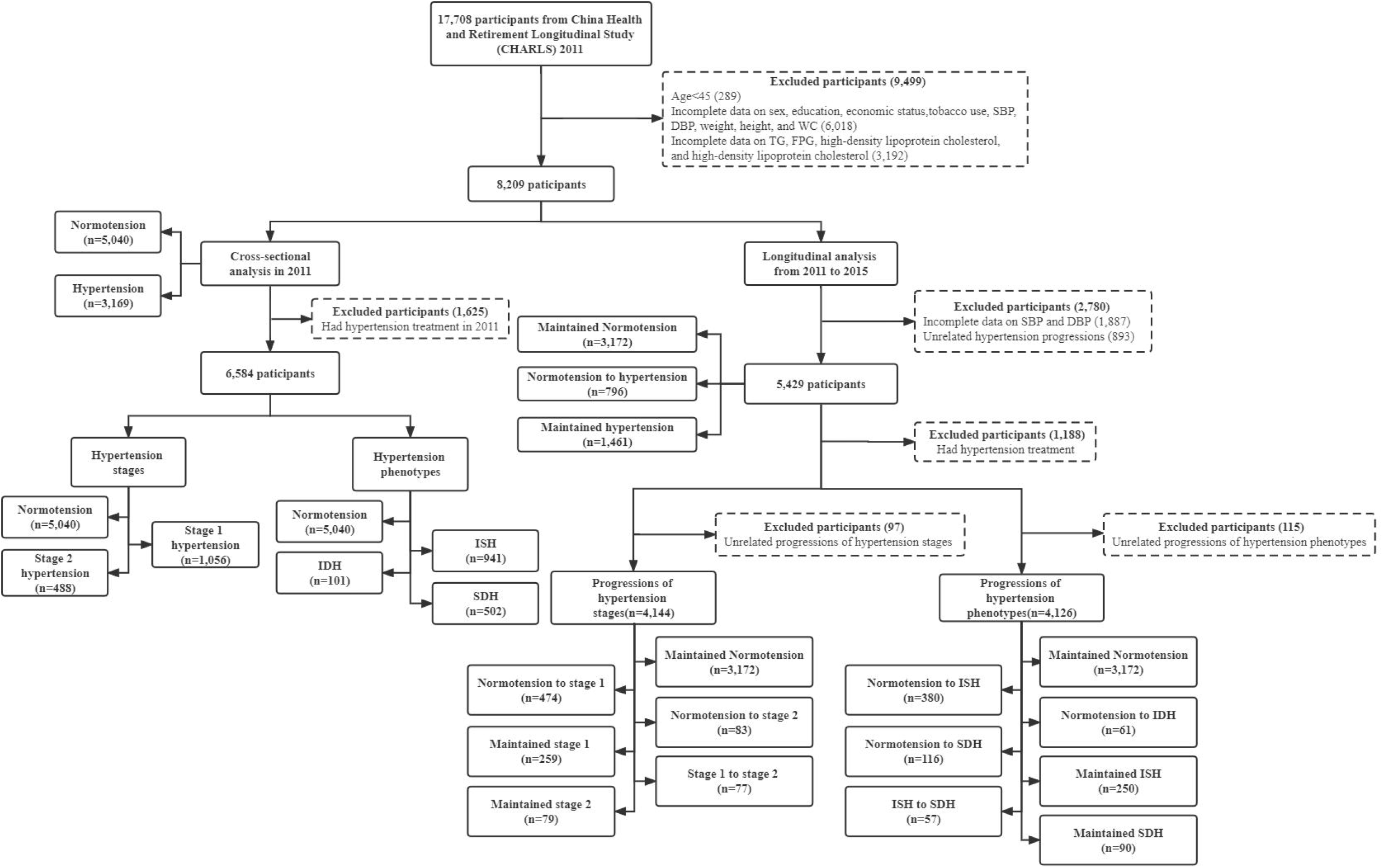
Flow chart of study participants Notes: SBP, systolic blood pressure. DBP, diastolic blood pressure. WC, waist circumstance. TG, fasting triglycerides. FPG, fasting plasma glucose. LDL-C, low-density lipoprotein cholesterol. HDL-C, high-density lipoprotein cholesterol. ISH, isolated systolic hypertension. IDH, isolated diastolic hypertension. SDH, systolic diastolic hypertension. Unrelated hypertension progressions: hypertension to normotension. Unrelated progressions of hypertension stages: stage 2 to stage 1, stage 2 to normotension, stage 1 to normotension. Unrelated progressions of hypertension phenotypes: ISH to normotension, ISH to IDH, IDH to normotension, IDH to ISH, SDH to normotension, SDH to ISH, SDH to IDH.

### Cross-sectional analyses

For the cross-sectional analysis in CHARLS 2011, we found that when compared with the bottom quartile of baseline TyG index, a higher quartile of TyG index was significantly associated with hypertension (odds ratio [OR] 1.17, 95% confidence interval [CI] 1.01-1.35 for quartile 2; OR 1.48, 95% CI 1.28-1.72 for quartile 3; OR 1.90, 95% CI 1.63-2.23 for quartile 4). Similar results were also found when the TyG index was used as a continuous variable and in the sex-stratified analyses (Table S4).

After excluding participants with antihypertensive medications in 2011 (n=1,625), baseline characteristics of the remaining 6,584 participants by hypertension stages and phenotypes were shown in Table S5 and Table S6, respectively. We also found that compared with people with the lowest quartile of baseline exposure, those with a higher TyG index had an increased risk of stage 1 hypertension (OR 1.26, 95% CI 1.02-1.55 for quartile 3; OR 1.71, 95% CI 1.38-2.13 for quartile 4) and stage 2 hypertension (OR 1.52, 95% CI 1.13-2.04 for quartile 3; OR 1.74, 95% CI 1.27-2.38 for quartile 4). Furthermore, the TyG index was also positively associated with ISH (OR 1.35, 95% CI 1.08-1.68 for quartile 3; OR 1.66, 95% CI 1.31-2.11 for quartile 4), IDH (OR 2.04, 95% CI 1.03-4.04 for quartile 3; OR 2.52, 95% CI 1.26-5.05 for quartile 4), and SDH (OR 1.65, 95% CI 1.23-2.23 for quartile 4) (Table 1). Similarly, a higher TyG index (per-unit) was significantly associated with the development of stage 1 hypertension (OR 1.43, 95% CI 1.27-1.61), stage 2 hypertension (OR 1.42, 95% CI 1.20-1.67), ISH (OR 1.41, 95% CI 1.24-1.61), IDH (OR 1.78, 95% CI 1.30-2.45), SDH (OR 1.35, 95% CI 1.15-1.58). However, neither as a continuous variable nor as a categorical variable of the TyG index, there was no evidence of a significant association between the TyG index and stage 2 hypertension in males.

**Table 1.** Association of the TyG index with hypertension stages and hypertension phenotypes in 2011

|  | Stage 1 hypertension | Stage 2 hypertension | ISH | IDH | SDH |
| --- | --- | --- | --- | --- | --- |
|  | Adjusted OR (95% CI) |  | Adjusted OR (95% CI) |  |  |
| Overall (N=6,584) |  |  |  |  |  |
| Quartile 1 | Reference | Reference | Reference | Reference | Reference |
| Quartile 2 | 1.10 (0.89-1.35) | 1.14 (0.85-1.54) | 1.10 (0.88-1.37) | 1.50 (0.75-3.00) | 1.07 (0.80-1.42) |
| Quartile 3 | 1.26 (1.02-1.55) | 1.52 (1.13-2.04) | 1.35 (1.08-1.68) | 2.04 (1.03-4.04) | 1.20 (0.90-1.62) |
| Quartile 4 | 1.71 (1.38-2.13) | 1.74 (1.27-2.38) | 1.66 (1.31-2.11) | 2.52 (1.26-5.05) | 1.65 (1.23-2.23) |
| TyG-continuous | 1.43 (1.27-1.61) | 1.42 (1.20-1.67) | 1.41 (1.24-1.61) | 1.78 (1.30-2.45) | 1.35 (1.15-1.58) |
| Male (N=3,154) |  |  |  |  |  |
| Quartile 1 | Reference | Reference | Reference | Reference | Reference |
| Quartile 2 | 1.08 (0.81-1.45) | 1.05 (0.70-1.58) | 1.06 (0.78-1.46) | 1.02 (0.41-2.55) | 1.11 (0.74-1.64) |
| Quartile 3 | 1.15 (0.85-1.55) | 1.24 (0.82-1.88) | 1.11 (0.80-1.54) | 1.16 (0.47-2.87) | 1.34 (0.91-1.99) |
| Quartile 4 | 1.74 (1.28-2.36) | 1.33 (0.85-2.09) | 1.65 (1.17-2.32) | 1.66 (0.68-4.04) | 1.62 (1.07-2.44) |
| TyG-continuous | 1.42 (1.20-1.67) | 1.25 (0.98-1.60) | 1.36 (1.12-1.65) | 1.65 (1.08-2.50) | 1.31 (1.06-1.62) |
| Female (N=3,430) |  |  |  |  |  |
| Quartile 1 | Reference | Reference | Reference | Reference | Reference |
| Quartile 2 | 1.17 (0.87-1.57) | 1.21 (0.79-1.87) | 1.22 (0.90-1.67) | 3.21 (1.01-10.20) | 0.91 (0.58-1.41) |
| Quartile 3 | 1.33 (0.98-1.79) | 1.74 (1.14-2.65) | 1.47 (1.08-2.01) | 4.40 (1.38-14.05) | 1.13 (0.73-1.76) |
| Quartile 4 | 1.79 (1.30-2.46) | 2.03 (1.30-3.19) | 1.81 (1.29-2.54) | 3.97 (1.17-13.43) | 1.74 (1.11-2.72) |
| TyG-continuous | 1.44 (1.21-1.71) | 1.45 (1.15-1.84) | 1.43 (1.19-1.72) | 1.73 (1.04-2.87) | 1.40 (1.10-1.78) |

Note: TyG, triglyceride-glucose. ISH, isolated systolic hypertension. IDH, isolated diastolic hypertension. SDH, systolic-diastolic hypertension. OR, odds ratio. CI, confidence interval. OR values were adjusted for age, sex, residence, education, economic status, tobacco use, alcohol consumption, general obesity, waist circumference, low-density lipoprotein cholesterol, high-density lipoprotein cholesterol, and C-reactive protein.

### Longitudinal analyses

In the longitudinal study using CHARLS 2011 and 2015, participants with incomplete data on SBP and DBP (n=1,887) or unrelated hypertension progressions (n=893) were further excluded, leaving 5,429 participants for longitudinal analyses. During the 4-year follow-up, 796 individuals (14.7%) developed hypertension (Table S7). Participants who were in the highest quartile TyG index were more likely to have the progression of normotension to hypertension (normotension to hypertension vs maintained normotension: OR 1.48, 95% CI 1.14-1.92) and maintained hypertension (maintained hypertension vs maintained normotension: OR 2.16, 95% CI 1.74-2.69), compared with those in the lowest quartile TyG index, as shown in Table S8. Associations of the per-unit TyG index with the progression of normotension to hypertension (OR 1.34, 95% CI 1.16-1.54) and maintained hypertension (OR 1.59, 95% CI 1.41-1.79) were also found when compared with maintained normotension. However, in females, there was no evidence of significant associations between the TyG index (per-unit) and normotension to hypertension (OR 1.18, 95% CI 0.96-1.45), compared with maintained normotension (Table S8).

Furthermore, 4,144 and 4,126 participants were enrolled separately to explore whether the TyG index was associated with the progressions of hypertension stages and phenotypes. Characteristics of these participants were described in Table S9 and Table S10, respectively. As shown in Table S11 and Figure 2, individuals with a higher baseline TyG index were more likely to have progressions of normotension to stage 1 (for highest quartile vs lowest quartile: OR 1.45, 95%CI 1.05-2.00; for per-unit: OR 1.39, 95% CI 1.16-1.65), maintained stage 1 (for highest quartile vs lowest quartile: OR 1.68, 95%CI 1.10-2.56; for per-unit: OR 1.32, 95% CI 1.05-1.67), and maintained stage 2 (for highest quartile vs lowest quartile: OR 2.62, 95%CI 1.23-5.62; for per-unit: OR 2.00, 95% CI 1.37-2.91).

**Figure 2.**
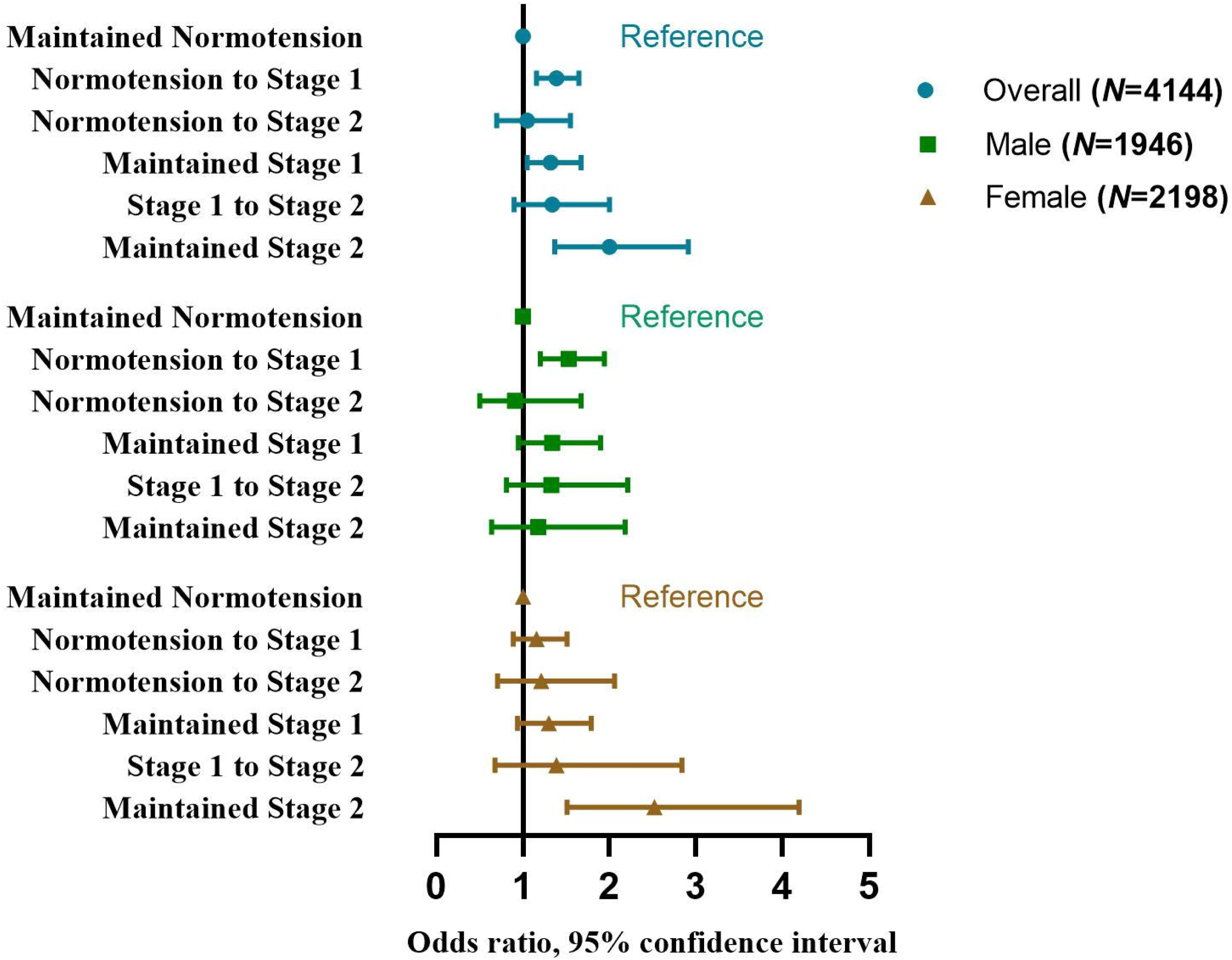
Odds ratios and 95% confidence interval of the progressions of hypertension stages by the TyG index

In addition, we also found significant associations of the TyG index with normotension to ISH (per-unit: OR 1.28, 95% CI 1.04-1.56), normotension to IDH (for highest quartile vs lowest quartile: OR 3.46, 95%CI 1.42-8.44; for per-unit: OR 1.94, 95% CI 1.27-2.97), maintained ISH (for highest quartile vs lowest quartile: OR 1.63, 95%CI 1.03-2.57; for per-unit: OR 1.34, 95% CI=1.05-1.70), and maintained SDH (for highest quartile vs lowest quartile: OR 2.66, 95%CI 1.36-5.20; for per-unit: OR 1.71, 95% CI 1.22-2.40) as shown in Table S12 and Figure 3.

**Figure 3.**
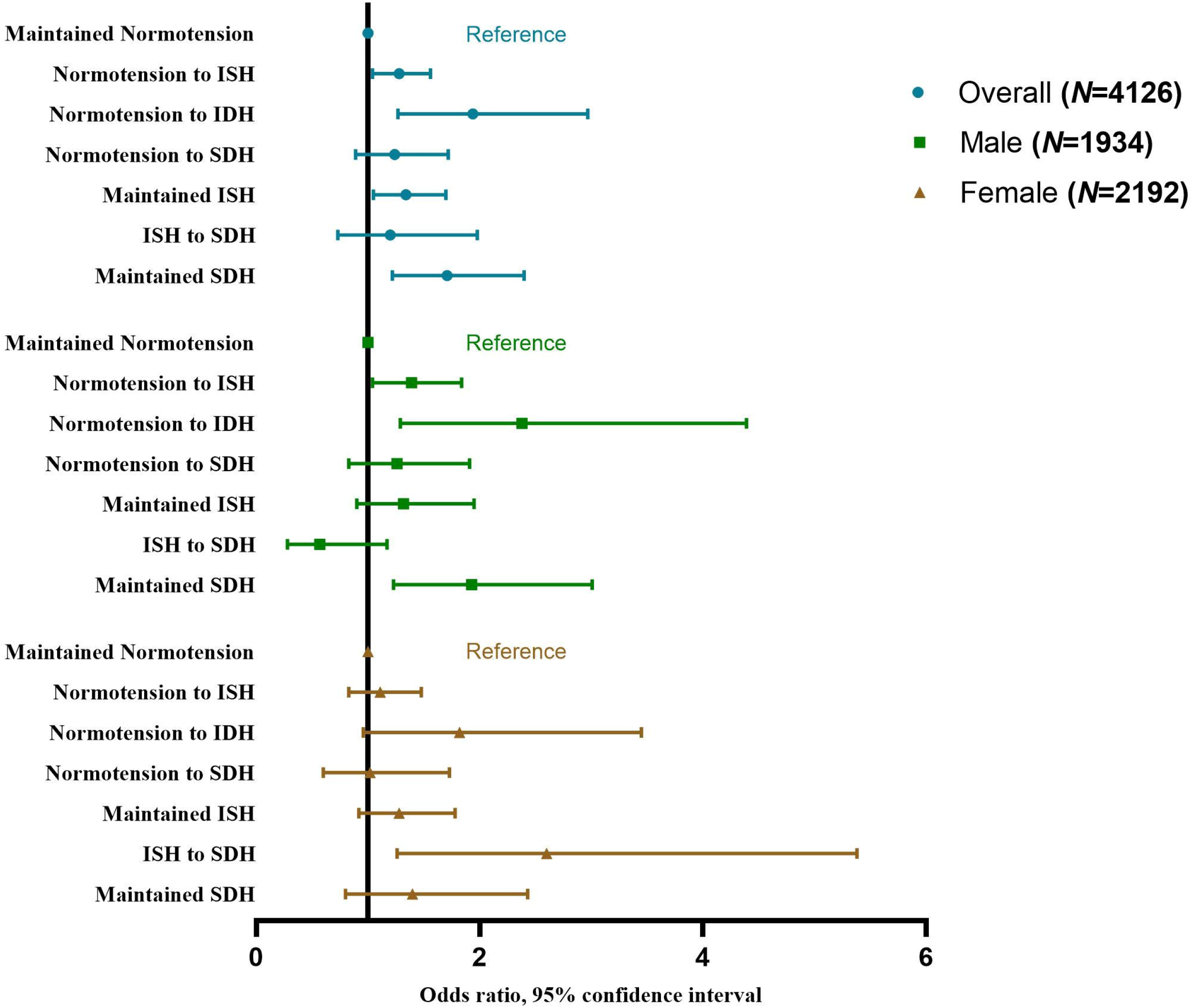
Odds ratios and 95% confidence interval of the progressions of hypertension phenotypes by the TyG index Notes: ISH, isolated systolic hypertension. IDH, isolated diastolic hypertension. SDH, systolic diastolic hypertension.

## Discussion

In this study, we found significant associations between a higher TyG index with increased risks of every hypertension stage and phenotype. Increased TyG index was also associated with the progressions of hypertension status (including normotension to hypertension and maintained hypertension), stages (including normotension to stage 1, maintained stage 1 and maintained stage 2) and phenotypes (including normotension to ISH, normotension to IDH, maintained ISH and maintained SDH).

Our results are consistent with previous findings, which discovered that a higher TyG index is associated with higher odds of hypertension^17,18,27-29^. Because of the TyG index’s high sensitivity and specificity, it has been proposed as a prominent indicator of IR^14,15^. And the association between IR and hypertension has been verified in some researches^30-32^. So, several potential mechanisms via IR may explain our main results. IR may cause hyperinsulinemia and elevate blood pressure by stimulating the sympathetic nervous system and the adrenergic system^33-35^. Increased insulin enhances insulin-mediated glucose metabolism in neurons and stimulates the sympathetic centre in the brain stem, resulting in higher BP^36^. What’s more, sympathetic excitement can generate increased catecholamine levels, which thicken vascular smooth muscle and lead to hypertension^37^. IR may also promote the release of angiotensin II by activating the renin-angiotensin system (RAS). Angiotensin II can inhibit the phosphatidylinositol 3-kinase pathway while activating the mitogen-activated protein kinase pathway and finally contribute to hypertension^35,38,39^. Meanwhile, elevated angiotensin II promotes adrenal aldosterone secretion and leads to sodium retention, plasma volume expansion, and non-genetic mineralocorticoid receptor mediation, all of which play an important role in hypertension pathogenesis^40,41^.

Our study supplemented previous findings and revealed that the TyG index was associated with multiple hypertension stages and phenotypes, as well as their progressions. Kaze et al.^42^ found a positive association between the development of IR and BP progressions, which aligns with our findings. In particular, we found that a higher TyG index was associated with minor hypertension progressions (i.e. normotension to stage 1, normotension to ISH, and normotension to IDH), implying that the TyG index could be a potential early screening indicator for hypertension. Given our sample limitations, we conducted our study in the general population but not in patients with hypertension, which may pull down the average TyG level and lead to the non-significant association of the TyG index with normotension to stage 2 and normotension to SDH. Therefore, the association of the TyG index with hypertension progressions across a broader range warrants further validation in larger-scale studies. However, in the sex-stratified analysis, we found that a higher TyG index was significantly associated with stage 2 hypertension in females, but not in males. The sex-stratified results may be related to the change in female estrogen levels. Research has shown that endogenous estrogen may play a role in higher insulin sensitivity in females^43^. Our study included females who were older than 45 years, and most of them were postmenopausal. Their dropping estrogen level resulted in a decrease of insulin sensitivity, making them susceptible to IR. Moreover, we found that a higher TyG index was significantly correlated with different progressions of hypertension in males (normotension to stage 1, normotension to ISH, normotension to IDH, maintained SDH), but not in females. Differences in the preferred location of fat storage between men and women may also affect the development of IR^44,45^. Women are more likely to store fat subcutaneously, while men store more abdominal fat with a higher amount of abdominal visceral fat and ectopic fat^46,47^. Therefore, the level of visceral fat storage will be higher in men than in women with the same BMI. Visceral fat is strongly associated with IR, which may lead to sex differences in the progression of hypertension.

## Strengths and limitations

The principal strength of this study is that we are the first to explore the associations of the TyG index with various hypertension stages and phenotypes, as well as their progressions, in a large Chinese general population. Since the TyG index is a recently proposed and high-profile indicator, our findings assist to accelerate the clinical application of the TyG index and facilitate early hypertension screening. Moreover, we used data from the CHARLS, which has broad geographic coverage, rigorous implementation procedures, and nationwide samples to warrant the generalizability of our conclusions.

Despite these strengths, several limitations need to be recognized. First, the TyG index was only assessed in 2011, and its changes during follow-up were not taken into account. Second, there were significant characteristic differences between included and excluded participants (Table S2), which may lead to selection bias and affect national representation. Third, since all participants were Chinese adults aged 45 and over, our findings cannot be directly extrapolated to younger groups. Fourth, we merged stage 2 and stage 3 hypertension into stage 2 hypertension due to the limited sample size, which may result in misjudging the association between the TyG index and stage 2 hypertension, or the progressions. Moreover, the participants in our study are the general population but not the patients with hypertension, which may pull down the average TyG level and lead to the non-significant association of the TyG index with normotension to stage 2 and normotension to SDH. More high-quality epidemiologic investigations and primary prevention studies are needed to explore the role of TyG index in the progression of hypertension. Finally, even though the model we used was adjusted for multiple covariates, certain possible confounding factors, such as exercise habits and family history of hypertension, were not included.

## Conclusion

In conclusion, our study revealed that a higher TyG index was significantly associated with hypertension stages, phenotypes, and their progressions. Given the rising prevalence of hypertension and diabetes in China, the TyG index may serve as a useful indicator for the early prevention of hypertension.

## Data Availability

The dataset presented in this study can be found in online repositories.

http://charls.pku.edu.cn/

## Ethics statement

The study protocol and data collection were approved by the Ethical Review Committee of Peking University. All respondents were informed and consented to the study protocol before data collection.

## Data availability statement

The dataset presented in this study can be found in online repositories. Requests to access the datasets should be directed to http://charls.pku.edu.cn/.

## Conflict of interest

The authors declare that the research was conducted in the absence of any commercial or financial relationships that could be construed as a potential conflict of interest.

## Funding

This study has no funding.

## Authors’ contributions

PS and DC designed the study. ZR and SS managed and analyzed the data. SS and KL prepared the first draft. PS, DC, SS, SL, KL, JC, WS, JZ, SZ and LH critically revised the manuscript. All authors were involved in revising the paper and gave final approval of the submitted versions.

## Acknowledgements

All authors thank the China Health and Retirement Longitudinal Study (CHARLS) team for providing data and instructions for using the dataset.

## Notes

### Competing Interest Statement

The authors have declared no competing interest.

### Funding Statement

This study did not receive any funding

