## supplementary table for "Associations of the Triglyceride and Glucose Index with Hypertension Stages, Phenotypes, and Progressions among Middle-aged and Older Chinese"

**Table Legends**

### Table S1. Classification of blood pressure in adults

| **Category** | | **Definition** |
| --- | --- | --- |
| **Normotension** | | SBP < 140 mmHg and DBP < 90 mmHg |
| **Hypertension** | | SBP ≥ 140 mmHg and/or DBP ≥ 90 mmHg and/or using antihypertensive medications |
| **Stages** | |  |
|  | Stage 1 | SBP = 140-159 mmHg and/or DBP = 90-99 mmHg |
|  | Stage 2 | SBP = 160-179 mmHg and/or DBP = 100-109 mmHg |
|  | Stage 3 | SBP ≥ 180 mmHg and/or DBP ≥ 110 mmHg |
| **Phenotypes** | |  |
|  | Isolated systolic hypertension | SBP ≥ 140 mmHg and DBP < 90 mmHg |
|  | Isolated diastolic hypertension | SBP < 140 mmHg and DBP ≥ 90 mmHg |
|  | Systolic diastolic hypertension | SBP ≥ 140 mmHg and DBP ≥ 90 mmHg |

Note: SBP, systolic blood pressure; DBP, diastolic blood pressure.

### Table S2. Characteristics of study participants and excluded participants

| **Characteristics** | | **Included Participants** | **Excluded participants** | ***P* value** |
| --- | --- | --- | --- | --- |
| No. of Participants | | 8,209 (46.4) | 9,499 (53.6) |  |
| Age, year | | 59.00 (52.00-65.00) | 57.00 (50.00-65.00) | 0.009 |
| Sex | |  |  | 0.068 |
|  | Male | 3,860 (47.0) | 4,611 (48.6) |  |
|  | Female | 4,349 (53.0) | 4,872 (51.4) |  |
| Residence | |  |  | <0.001 |
|  | Urban | 2,816 (34.3) | 4,352 (45.8) |  |
|  | Rural | 5,393 (65.7) | 5,144 (54.2) |  |
| Education | |  |  | <0.001 |
|  | Illiterate | 2,345 (28.6) | 2,458 (26.0) |  |
|  | Primary school and below | 3,410 (41.5) | 3,542 (37.5) |  |
|  | Junior high school or above | 2,454 (29.9) | 3,444 (36.5) |  |
| Economic status | |  |  | <0.001 |
|  | Poor | 2,741 (33.4) | 1,854 (27.3) |  |
|  | Middle | 2,803 (34.2) | 2,085 (30.7) |  |
|  | Rich | 2,665 (32.5) | 2,856 (42.0) |  |
| Tobacco use | |  |  | 0.757 |
|  | Never smoking | 4,958 (60.4) | 5,668 (60.6) |  |
|  | Ever smoking | 3,251 (39.6) | 3,680 (39.4) |  |
| Alcohol consumption | |  |  | 0.330 |
|  | Never drinking | 6,131 (74.7) | 7,036 (75.3) |  |
|  | Ever drinking | 2,078 (25.3) | 2,305 (24.7) |  |
| Hypertension status | |  |  | <0.001 |
|  | Normotension | 5,040 (61.4) | 6,256 (65.9) |  |
|  | Hypertension | 3,169 (38.6) | 3,240 (34.1) |  |
| General obesity | |  |  | 0.034 |
|  | Normal | 4,816 (58.7) | 3,319 (61.1) |  |
|  | Overweight | 954 (11.6) | 613 (11.3) |  |
|  | Obesity | 2,439 (29.7) | 1,499 (27.6) |  |
| WC, cm | | 84.70 (78.00-92.00) | 84.00 (77.00-91.60) | 0.010 |
| LDL-C, mg/dL | | 114.43 (93.17-137.24) | 112.89 (91.24-135.70) | 0.008 |
| HDL-C, mg/dL | | 49.48 (40.21-59.92) | 48.33 (39.43-58.38) | <0.001 |
| CRP, mg/L | | 1.04 (0.55-2.19) | 1.04 (0.54-2.02) | 0.599 |
| The TyG index | | 8.60 (8.22-9.05) | 8.62 (8.25-9.10) | 0.031 |

Note: WC, waist circumstance. LDL-C, low-density lipoprotein cholesterol. HDL-C, high-density lipoprotein cholesterol. CRP, C-reactive protein. The TyG index, triglyceride-glucose index. Values are presented as number (N) with percent (%) for categorical variables or median with interquartile range (IQR) for continuous variables. Excluded participants have missing data on age, sex, education, economic status, tobacco use, alcohol consumption, general obesity, waist circumstance, low-density lipoprotein cholesterol, high-density lipoprotein cholesterol, and C-reactive protein.

### Table S3. Characteristics of study participants by hypertension statuses in 2011 (*N*=8,209)

| **Characteristics** | | **Normotension** | **Hypertension** | ***P* value** |
| --- | --- | --- | --- | --- |
| No. of Participants | | 5,040 (61.4) | 3,169 (38.6) |  |
| Age, year | | 57.00 (50.00-63.00) | 61.00 (55.00-69.00) | <0.001 |
| Sex | |  |  | 0.068 |
|  | Male | 2,410 (47.8) | 1,450 (45.8) |  |
|  | Female | 2,630 (52.2) | 1,719 (54.2) |  |
| Residence | |  |  | <0.001 |
|  | Urban | 1,634 (32.4) | 1,182 (37.3) |  |
|  | Rural | 3,406 (67.6) | 1,987 (62.7) |  |
| Education | |  |  | <0.001 |
|  | Illiterate | 1,326 (26.3) | 1,019 (32.2) |  |
|  | Primary school and below | 2,092 (41.5) | 1,318 (41.6) |  |
|  | Junior high school or above | 1,622 (32.2) | 832 (26.3) |  |
| Economic status | |  |  | 0.019 |
|  | Poor | 1,650 (32.7) | 1,091 (34.4) |  |
|  | Middle | 1,696 (33.7) | 1,107 (34.9) |  |
|  | Rich | 1,694 (33.6) | 971 (30.6) |  |
| Tobacco use | |  |  | 0.095 |
|  | Never smoking | 3,008 (59.7) | 1,950 (61.5) |  |
|  | Ever smoking | 2,032 (40.3) | 1,219 (38.5) |  |
| Alcohol consumption | |  |  | 0.032 |
|  | Never drinking | 3,723 (73.9) | 2,408 (76.0) |  |
|  | Ever drinking | 1,317 (26.1) | 761 (24.0) |  |
| General obesity | |  |  | <0.001 |
|  | Normal | 3,308 (65.6) | 1,508 (47.6) |  |
|  | Overweight | 1,350 (26.8) | 1,089 (34.4) |  |
|  | Obesity | 382 (7.6) | 572 (18.1) |  |
| WC, cm | | 82.80 (76.40-89.40) | 88.20 (81.00-95.80) | <0.001 |
| LDL-C, mg/dL | | 113.27 (92.78-134.92) | 116.37 (94.72-141.50) | <0.001 |
| HDL-C, mg/dL | | 50.64 (41.37-60.70) | 47.55 (39.05-57.99) | <0.001 |
| CRP, mg/L | | 0.90 (0.50-1.90) | 1.29 (0.66-2.69) | <0.001 |
| The TyG index | | 8.52 (8.17-8.94) | 8.74 (8.34-9.20) | <0.001 |

Note: WC, waist circumstance. LDL-C, low-density lipoprotein cholesterol. HDL-C, high-density lipoprotein cholesterol. CRP, C-reactive protein. The TyG index, triglyceride-glucose index. Values are presented as number (N) with percent (%) for categorical variables or median with interquartile range (IQR) for continuous variables.

### Table S4. Association between TyG index and hypertension in 2011

| **Variables** | **Overall (N=8,209)** | **Male (N=3,860)** | **Female (N=4,349)** |
| --- | --- | --- | --- |
|  | **Adjusted OR (95% CI)** | | |
| Quartile 1 | Reference | Reference | Reference |
| Quartile 2 | 1.17 (1.01-1.35) | 1.07 (0.88-1.32) | 1.17 (0.96-1.42) |
| Quartile 3 | 1.48 (1.28-1.72) | 1.33 (1.08-1.64) | 1.57 (1.28-1.93) |
| Quartile 4 | 1.90 (1.63-2.23) | 1.64 (1.31-2.05) | 1.99 (1.59-2.48) |
| TyG-continuous | 1.45 (1.33-1.58) | 1.38 (1.23-1.56) | 1.48 (1.31-1.67) |

Note: TyG, triglyceride-glucose. OR, odds ratio. CI, confidence interval. OR values were adjusted for age, sex, residence, education, economic status, tobacco use, alcohol consumption, body mass index, waist circumstance, low-density lipoprotein cholesterol, high-density lipoprotein cholesterol, and C-reactive protein.

### Table S5. Characteristics of study participants by hypertension stages in 2011 (*N*=6,584)

| **Characteristics** | | **Normotension** | **Stage 1 hypertension** | **Stage 2 hypertension** | ***P* value** |
| --- | --- | --- | --- | --- | --- |
| No. of Participants | | 5,040 (76.6) | 1,056 (16.0) | 488 (7.4) |  |
| Age, year | | 57.00 (50.00-63.00) | 61.00 (54.00-69.00) | 61.50 (56.00-70.00) | <0.001 |
| Sex | |  |  |  | 0.399 |
|  | Male | 2,410 (47.8) | 521 (49.3) | 223 (45.7) |  |
|  | Female | 2,630 (52.2) | 535 (50.6) | 265 (54.3) |  |
| Residence | |  |  |  | 0.303 |
|  | Urban | 1,634 (32.4) | 366 (34.7) | 167 (34.2) |  |
|  | Rural | 3,406 (67.6) | 690 (65.3) | 321 (65.8) |  |
| Education | |  |  |  | <0.001 |
|  | Illiterate | 1,326 (26.3) | 354 (33.5) | 192 (39.3) |  |
|  | Primary school and below | 2,092 (41.5) | 428 (40.5) | 203 (41.6) |  |
|  | Junior high school or above | 1,622 (32.2) | 274 (26.0) | 93 (19.1) |  |
| Economic status | |  |  |  | <0.001 |
|  | Poor | 1,650 (32.7) | 378 (35.8) | 195 (40.0) |  |
|  | Middle | 1,696 (33.7) | 376 (35.6) | 157 (32.2) |  |
|  | Rich | 1,694 (33.6) | 302 (28.6) | 136 (27.9) |  |
| Tobacco use | |  |  |  | 0.219 |
|  | Never smoking | 3,008 (59.7) | 603 (57.1) | 298 (61.1) |  |
|  | Ever smoking | 2,032 (40.3) | 453 (42.9) | 190 (38.9) |  |
| Alcohol consumption | |  |  |  | 0.304 |
|  | Never drinking | 3,723 (73.9) | 760 (72.0) | 350 (71.7) |  |
|  | Ever drinking | 1,317 (26 1) | 296 (28.0) | 138 (28.3) |  |
| Systolic blood pressure, mmHg | | 118.67 (110.00-127.67) | 146.00 (142.00-150.67) | 171.00 (163.67-181.67) | <0.001 |
| Diastolic blood pressure, mmHg | | 70.67 (64.33-77.00) | 84.33 (77.67-90.67) | 93.00 (83.50-101.00) | <0.001 |
| General obesity | |  |  |  | <0.001 |
|  | Normal | 3,308 (65.6) | 613 (58.1) | 283 (58.0) |  |
|  | Overweight | 1,350 (26.8) | 322 (30.5) | 145 (29.7) |  |
|  | Obesity | 382 (7.6) | 121 (11.5) | 60 (12.3) |  |
| WC, cm | | 82.80(76.40-89.40) | 86.00 (78.60-93.00) | 87.0 (79.20-93.40) | <0.001 |
| LDL-C, mg/dL | | 113.27 (92.78-134.92) | 115.59 (94.33-138.02) | 118.49 (94.52-139.37) | 0.017 |
| HDL-C, mg/dL | | 50.64 (41.37-60.70) | 49.87 (40.98-61.08) | 50.26 (40.98-60.12) | 0.779 |
| CRP, mg/L | | 0.90 (0.50-1.90) | 1.14 (0.60-2.38) | 1.16 (0.60-2.51) | <0.001 |
| The TyG index | | 8.52 (8.17-8.94) | 8.66 (8.27-9.13) | 8.68 (8.30-9.08) | <0.001 |

Note: WC, waist circumstance. LDL-C, low-density lipoprotein cholesterol. HDL-C, high-density lipoprotein cholesterol. CRP, C-reactive protein. The TyG index, triglyceride-glucose index. Values are presented as number (N) with percent (%) for categorical variables or median with interquartile range (IQR) for continuous variables.

### Table S6. Characteristics of study participants by hypertension phenotypes in 2011 (*N*=6,584)

| **Characteristics** | | **Normotension** | **ISH** | **IDH** | **SDH** | ***P* value** |
| --- | --- | --- | --- | --- | --- | --- |
| No. of Participants | | 5,040 (76.6) | 941 (14.3) | 101 (1.5) | 502 (7.6) |  |
| Age, year | | 57.00 (50.00-63.00) | 64.00 (58.00-72.00) | 52.00 (48.00-56.00) | 57.00 (51.00-62.00) | <0.001 |
| Sex | |  |  |  |  | <0.001 |
|  | Male | 2,410 (47.8) | 413 (43.9) | 56 (55.5) | 275 (54.8) |  |
|  | Female | 2,630 (52.2) | 528 (56.1) | 45 (44.6) | 227 (45.2) |  |
| Residence | |  |  |  |  | 0.011 |
|  | Urban | 1,634 (32.4) | 306 (32.5) | 47 (46.5) | 180 (35.9) |  |
|  | Rural | 3,406 (67.6) | 635 (67.5) | 54 (53.5) | 322 (64.1) |  |
| Education | |  |  |  |  | <0.001 |
|  | Illiterate | 1,326 (26.3) | 400 (42.5) | 16 (15.8) | 130 (25.9) |  |
|  | Primary school and below | 2,092 (41.5) | 382 (40.6) | 30 (29.7) | 219 (43.6) |  |
|  | Junior high school or above | 1,622 (32.2) | 159 (16.9) | 55 (54.5) | 153 (30.5) |  |
| Economic status | |  |  |  |  | <0.001 |
|  | Poor | 1,650 (32.7) | 367 (39.0) | 23 (22.8) | 183 (36.5) |  |
|  | Middle | 1,696 (33.7) | 327 (34.8) | 32 (31.7) | 174 (34.7) |  |
|  | Rich | 1,694 (33.6) | 247 (26.3) | 46 (45.5) | 145 (28.9) |  |
| Tobacco use | |  |  |  |  | 0.034 |
|  | Never smoking | 3,008 (59.7) | 575 (61.1) | 57 (56.4) | 269 (53.6) |  |
|  | Ever smoking | 2,032 (40.3) | 366 (38.9) | 44 (43.6) | 233 (46.4) |  |
| Alcohol consumption | |  |  |  |  | <0.001 |
|  | Never drinking | 3,723 (73.9) | 719 (76.4) | 57 (56.4) | 334 (66.5) |  |
|  | Ever drinking | 1,317 (26 1) | 222 (23.6) | 44 (43.6) | 168 (33.5) |  |
| Systolic blood pressure, mmHg | | 118.67 (110.00-127.67) | 148.00 (143.67-157.0) | 134.00 (129.33-137.33) | 159.33 (149.33-175.33) | <0.001 |
| Diastolic blood pressure, mmHg | | 70.67 (64.33-77.00) | 80.67 (75.00-85.33) | 92.67 (91.00-95.00) | 96.00 (92.67-100.67) | <0.001 |
| General obesity | |  |  |  |  | <0.001 |
|  | Normal | 3,308 (65.6) | 602 (64.0) | 45 (44.6) | 249 (49.6) |  |
|  | Overweight | 1,350 (26.8) | 257 (27.3) | 47 (46.5) | 163 (32.5) |  |
|  | Obesity | 382 (7.6) | 82 (8.7) | 9 (8.9) | 90 (17.9) |  |
| WC, cm | | 82.80(76.40-89.40) | 85.00 (78.00-92.00) | 88.00 (80.20-94.40) | 87.95 (80.00-94.90) | <0.001 |
| LDL-C, mg/dL | | 113.27 (92.78-134.92) | 117.53 (95.88-140.72) | 122.55 (91.24-144.20) | 113.47 (92.40-134.92) | 0.002 |
| HDL-C, mg/dL | | 50.64 (41.37-60.70) | 50.26 (42.14-61.86) | 49.10 (37.11-55.67) | 48.33 (40.21-59.54) | 0.009 |
| CRP, mg/L | | 0.90 (0.50-1.90) | 1.12 (0.58-2.31) | 1.08 (0.65-3.04) | 1.23 (0.61-2.59) | <0.001 |
| The TyG index | | 8.52 (8.17-8.94) | 8.65 (8.26-9.07) | 8.76 (8.39-9.34) | 8.68 (8.28-9.15) | <0.001 |

Note: WC, waist circumstance. LDL-C, low-density lipoprotein cholesterol. HDL-C, high-density lipoprotein cholesterol. CRP, C-reactive protein. The TyG index, triglyceride-glucose index. Values are presented as number (N) with percent (%) for categorical variables or median with interquartile range (IQR) for continuous variables.

### Table S7. Characteristics of study participants by progressions of hypertension statuses from 2011 to 2015 (*N*=5,429)

| **Characteristics** | | **Maintained Normotension** | **Normotension to hypertension** | **Maintained hypertension** | ***P* value** |
| --- | --- | --- | --- | --- | --- |
| No. of Participants | | 3,172 (58.4) | 796 (14.7) | 1,461 (26.9) |  |
| Age, year | | 56.00 (50.00-62.00) | 59.00 (52.00-66.00) | 61.00 (55.00-68.00) | <0.001 |
| Sex | |  |  |  | 0.153 |
|  | Male | 1,460 (46.0) | 395 (49.6) | 668 (45.7) |  |
|  | Female | 1,712 (54.0) | 401 (50.4) | 793 (54.3) |  |
| Residence | |  |  |  | 0.008 |
|  | Urban | 944 (29.8) | 244 (30.7) | 501 (34.3) |  |
|  | Rural | 2,228 (70.2) | 552 (69.4) | 960 (65.7) |  |
| Education | |  |  |  | <0.001 |
|  | Illiterate | 820 (25.9) | 237 (29.8) | 464 (31.8) |  |
|  | Primary school and below | 1,342 (42.3) | 339 (42.6) | 615 (42.1) |  |
|  | Junior high school or above | 1,010 (31.8) | 220 (27.6) | 382 (26.2) |  |
| Economic status | |  |  |  | 0.136 |
|  | Poor | 1,053 (33.2) | 294 (36.9) | 518 (35.5) |  |
|  | Middle | 1,084 (34.2) | 269 (33.8) | 505 (34.6) |  |
|  | Rich | 1,035 (32.6) | 233 (29.3) | 438 (30.0) |  |
| Tobacco use | |  |  |  | 0.807 |
|  | Never smoking | 1,937 (61.1) | 477 (59.9) | 895 (61.3) |  |
|  | Ever smoking | 1,235 (38.9) | 319 (40.1) | 566 (38.7) |  |
| Alcohol consumption | |  |  |  | 0.015 |
|  | Never drinking | 2,384 (75.2) | 559 (70.2) | 1,095 (75.0) |  |
|  | Ever drinking | 788 (24.8) | 237 (29.8) | 366 (25.1) |  |
| General obesity | |  |  |  | <0.001 |
|  | Normal | 2,150 (67.8) | 465 (58.4) | 662 (45.3) |  |
|  | Overweight | 795 (25.1) | 250 (31.4) | 538 (36.8) |  |
|  | Obesity | 227 (7.2) | 81 (10.2) | 261 (17.9) |  |
| WC, cm | | 82.10 (76.00-89.00) | 84.35 (78.00-92.00) | 88.70 (81.20-96.00) | <0.001 |
| LDL-C, mg/dL | | 112.50 (92.40-134.15) | 115.59 (95.10-138.60) | 116.75 (94.33-141.50) | <0.001 |
| HDL-C mg/dL | | 50.64 (41.37-60.70) | 50.26 (40.59-61.08) | 47.55 (39.05-57.99) | <0.001 |
| CRP, mg/L | | 0.88 (0.50-1.85) | 0.91 (0.51-1.78) | 1.28 (0.65-2.63) | <0.001 |
| The TyG index | | 8.51 (8.16-8.91) | 8.60 (8.23-9.01) | 8.75 (8.35-9.19) | <0.001 |

Note: WC, waist circumstance. LDL-C, low-density lipoprotein cholesterol. HDL-C, high-density lipoprotein cholesterol. CRP, C-reactive protein. The TyG index, triglyceride-glucose index. Values are presented as number (N) with percent (%) for categorical variables or median with interquartile range (IQR) for continuous variables.

### Table S8. Association between TyG index in 2011 and the progressions of hypertension statuses from 2011 to 2015

|  |  | **Normotension to hypertension** | **Maintained hypertension** |
| --- | --- | --- | --- |
|  |  | **Adjusted OR (95% CI)** | |
| ***Overall (N=5,429)*** | | | |
|  | Quartile 1 | Reference | Reference |
|  | Quartile 2 | 1.14 (0.92-1.43) | 1.19 (0.97-1.45) |
|  | Quartile 3 | 1.32 (1.04-1.67) | 1.63 (1.33-1.99) |
|  | Quartile 4 | 1.48 (1.14-1.92) | 2.16 (1.74-2.69) |
|  | TyG-continuous | 1.34 (1.16-1.54) | 1.59 (1.41-1.79) |
| ***Male (N=2,523)*** | | | |
|  | Quartile 1 | Reference | Reference |
|  | Quartile 2 | 0.84 (0.60-1.16) | 1.01 (0.75-1.35) |
|  | Quartile 3 | 1.37 (0.98-1.90) | 1.51 (1.12-2.02) |
|  | Quartile 4 | 1.47 (1.03-2.11) | 1.67 (1.22-2.30) |
|  | TyG-continuous | 1.50 (1.21-1.85) | 1.54 (1.29-1.83) |
| ***Female (N=2,906)*** | | | |
|  | Quartile 1 | Reference | Reference |
|  | Quartile 2 | 1.24 (0.90-1.71) | 1.12 (0.85-1.48) |
|  | Quartile 3 | 1.44 (1.03-2.01) | 1.67 (1.26-2.22) |
|  | Quartile 4 | 1.45 (1.00-2.11) | 2.33 (1.71-3.17) |
|  | TyG-continuous | 1.18 (0.96-1.45) | 1.64 (1.38-1.94) |

Note: TyG, triglyceride-glucose. OR, odds ratio. CI, confidence interval. OR values were adjusted for age, sex, residence, education, economic status, tobacco use, alcohol consumption, general obesity, waist circumstance, low-density lipoprotein cholesterol, high-density lipoprotein cholesterol, and C-reactive protein.

### Table S9. Characteristics of study participants by the progressions of hypertension stages from 2011 to 2015 (*N*=4,144)

| **Characteristics** | | **Maintained Normotension** | **Normotension to stage 1** | **Normotension to stage 2** | **Maintained stage 1** | **Stage 1 to Stage 2** | **Maintained stage 2** | ***P* value** |
| --- | --- | --- | --- | --- | --- | --- | --- | --- |
| No. of Participants | | 3,172 (76.5) | 474 (11.4) | 83 (2.0) | 259 (6.3) | 77 (1.9) | 79 (1.0) |  |
| Age, year | | 56.00 (50.00-62.00) | 59.00 (52.00-66.00) | 64.00 (54.00-68.00) | 60.00 (54.00-69.00) | 63.00 (54.00-69.00) | 61.00 (57.00-68.00) | <0.001 |
| Sex | |  |  |  |  |  |  | 0.005 |
|  | Male | 1,460 (46.0) | 249 (52.5) | 38 (45.8) | 115 (44.4) | 49 (63.6) | 35 (44.3) |  |
|  | Female | 1,712 (54.0) | 225 (47.5) | 45 (54.2) | 144 (55.6) | 28 (36.4) | 44 (55.7) |  |
| Residence | |  |  |  |  |  |  | 0.010 |
|  | Urban | 944 (29.8) | 157 (33.1) | 20 (24.1) | 86 (33.2) | 29 (37.7) | 36 (45.6) |  |
|  | Rural | 2,228 (70.2) | 317 (66.9) | 63 (75.9) | 173 (66.8) | 48 (62.3) | 43 (54.4) |  |
| Education | |  |  |  |  |  |  | <0.001 |
|  | Illiterate | 820 (25.9) | 147 (31.0) | 26 (31.3) | 89 (34.4) | 26 (33.8) | 31 (39.2) |  |
|  | Primary school and below | 1,342 (42.3) | 184 (38.8) | 42 (50.6) | 109 (42.1) | 27 (35.1) | 32 (40.5) |  |
|  | Junior high school or above | 1,010 (31.8) | 143 (30.2) | 15 (18.1) | 61 (23.6) | 24 (31.2) | 16 (20.3) |  |
| Economic status | |  |  |  |  |  |  | 0.024 |
|  | Poor | 1,053 (33.2) | 192 (40.5) | 28 (33.7) | 107 (41.3) | 22 (28.6) | 34 (43.0) |  |
|  | Middle | 1,084 (34.2) | 149 (31.4) | 32 (38.6) | 79 (30.5) | 29 (37.7) | 21 (26.6) |  |
|  | Rich | 1,035 (32.6) | 133 (28.1) | 23 (27.7) | 73 (28.2) | 26 (33.8) | 24 (30.4) |  |
| Tobacco use | |  |  |  |  |  |  | 0.148 |
|  | Never smoking | 1,937 (61.1) | 275 (58.0) | 53 (63.9) | 157 (60.6) | 36 (46.8) | 47 (59.5) |  |
|  | Ever smoking | 1,235 (38.9) | 199 (42.0) | 30 (36.1) | 102 (39.4) | 41 (53.3) | 32 (40.5) |  |
| Alcohol consumption | |  |  |  |  |  |  | <0.001 |
|  | Never drinking | 2,384 (75.2) | 331 (69.8) | 63 (75.9) | 194 (74.9) | 41 (53.3) | 57 (72.2) |  |
|  | Ever drinking | 788 (24.8) | 143 (30.2) | 20 (24.1) | 65 (25.1) | 36 (46.8) | 22 (27.9) |  |
| General obesity | |  |  |  |  |  |  | <0.001 |
|  | Normal | 2,150 (67.8) | 285 (60.1) | 57 (68.7) | 141 (54.4) | 48 (62.3) | 44 (55.7) |  |
|  | Overweight | 795 (25.1) | 143 (30.2) | 19 (22.9) | 86 (33.2) | 21 (27.3) | 27 (34.2) |  |
|  | Obesity | 227 (7.2) | 46 (9.7) | 7 (8.4) | 32 (12.4) | 8 (10.4) | 8 (10.1) |  |
| WC, cm | | 82.10 (76.00-89.00) | 84.10 (77.40-91.40) | 83.00 (78.00-91.00) | 87.00 (79.20-93.60) | 87.20 (81.40-91.00) | 88.00 (80.00-93.50) | <0.001 |
| LDL-C, mg/dL | | 112.50 (92.40-134.15) | 115.59 (95.10-140.72) | 111.73 (93.94-129.51) | 119.07 (95.49-143.04) | 110.95 (84.67-131.06) | 117.53 (86.99-144.98) | 0.030 |
| HDL-C, mg/dL | | 50.64 (41.37-60.70) | 50.26 (40.59-61.47) | 50.64 (40.98-61.86) | 49.10 (41.37-58.76) | 49.48 (42.53-62.63) | 48.33 (37.11-60.70) | 0.585 |
| CRP, mg/L | | 0.88 (0.50-1.85) | 0.92 (0.52-1.78) | 0.88 (0.46-1.78) | 1.16 (0.59-2.42) | 0.95 (0.56-1.87) | 1.27 (0.62-2.29) | 0.005 |
| The TyG index | | 8.51 (8.16-8.91) | 8.60 (8.23-9.00) | 8.50 (8.22-8.90) | 8.62 (8.25-9.12) | 8.60 (8.23-9.08) | 8.69 (8.45-9.17) | <0.001 |

Note: WC, waist circumstance. LDL-C, low-density lipoprotein cholesterol. HDL-C, high-density lipoprotein cholesterol. CRP, C-reactive protein. The TyG index, triglyceride-glucose index. Values are presented as number (N) with percent (%) for categorical variables or median with interquartile range (IQR) for continuous variables.

### Table S10. Characteristics of study participants by the progressions of hypertension phenotypes from 2011 to 2015 (*N*=4,126)

| **Characteristics** | | **Maintained Normotension** | **Normotension to ISH** | **Normotension to IDH** | **Normotension to SDH** | **Maintained ISH** | **ISH to SDH** | **Maintained SDH** | ***P* value** |
| --- | --- | --- | --- | --- | --- | --- | --- | --- | --- |
| No. of Participants | | 3,172 (76.9) | 380 (9.2) | 61 (1.5) | 116 (2.8) | 250 (6.1) | 57 (1.4) | 90 (2.2) |  |
| Age, year | | 56.00 (50.00-62.00) | 62.00 (56.00-68.00) | 51.00 (47.00-57.00) | 55.00 (49.00-61.00) | 66.00 (59.00-72.00) | 61.00 (56.00-66.00) | 56.00 (49.00-60.00) | <0.001 |
| Sex | |  |  |  |  |  |  |  | 0.007 |
|  | Male | 1,460 (46.0) | 192 (50.5) | 30 (49.2) | 65 (56.0) | 102 (40.8) | 33 (57.9) | 52 (57.8) |  |
|  | Female | 1,712 (54.0) | 188 (49.5) | 31 (50.8) | 51 (44.0) | 148 (59.2) | 24 (42.1) | 38 (42.2) |  |
| Residence | |  |  |  |  |  |  |  | 0.030 |
|  | Urban | 944 (29.8) | 126 (33.2) | 18 (29.5) | 33 (28.5) | 89 (35.6) | 20 (35.1) | 40 (44.4) |  |
|  | Rural | 2,228 (70.2) | 254 (66.8) | 43 (70.5) | 83 (71.6) | 161 (64.4) | 37 (64.9) | 50 (55.6) |  |
| Education | |  |  |  |  |  |  |  | <0.001 |
|  | Illiterate | 820 (25.9) | 134 (35.3) | 11 (18.0) | 28 (24.1) | 112 (44.8) | 18 (31.6) | 18 (20.0) |  |
|  | Primary school and below | 1,342 (42.3) | 155 (40.8) | 23 (37.7) | 48 (41.4) | 104 (41.6) | 21 (36.8) | 40 (44.4) |  |
|  | Junior high school or above | 1,010 (31.8) | 91 (24.0) | 27 (44.3) | 40 (34.5) | 34 (13.6) | 18 (31.6) | 32 (35.6) |  |
| Economic status | |  |  |  |  |  |  |  | 0.005 |
|  | Poor | 1,053 (33.2) | 162 (42.6) | 20 (32.8) | 38 (32.8) | 104 (41.6) | 21 (36.8) | 34 (37.8) |  |
|  | Middle | 1,084 (34.2) | 126 (33.2) | 14 (23.0) | 41 (35.3) | 73 (29.2) | 20 (35.1) | 29 (32.2) |  |
|  | Rich | 1,035 (32.6) | 92 (24.2) | 27 (44.3) | 37 (31.9) | 73 (29.2) | 16 (28.1) | 27 (30.0) |  |
| Tobacco use | |  |  |  |  |  |  |  | 0.045 |
|  | Never smoking | 1,937 (61.1) | 223 (58.7) | 39 (63.9) | 66 (56.9) | 161 (64.4) | 26 (45.6) | 45 (50.0) |  |
|  | Ever smoking | 1,235 (38.9) | 157 (41.3) | 22 (36.1) | 50 (43.1) | 89 (35.6) | 31 (54.4) | 45 (50.0) |  |
| Alcohol consumption | |  |  |  |  |  |  |  | 0.008 |
|  | Never drinking | 2,384 (75.2) | 272 (71.6) | 44 (72.1) | 78 (67.2) | 192 (76.8) | 39 (68.4) | 54 (60.0) |  |
|  | Ever drinking | 788 (24.8) | 108 (28.4) | 17 (27.9) | 38 (32.8) | 58 (23.2) | 18 (31.6) | 36 (40.0) |  |
| General obesity | |  |  |  |  |  |  |  | <0.001 |
|  | Normal | 2,150 (67.8) | 244 (64.2) | 36 (59.0) | 62 (53.5) | 153 (61.2) | 32 (56.1) | 44 (48.9) |  |
|  | Overweight | 227 (7.2) | 30 (7.9) | 7 (11.5) | 16 (13.8) | 26 (10.4) | 4 (7.0) | 14 (15.6) |  |
|  | Obesity | 795 (25.1) | 106 (27.9) | 18 (29.5) | 38 (32.8) | 71 (28.4) | 21 (36.8) | 32 (35.6) |  |
| WC, cm | | 82.10 (76.00-89.00) | 83.30 (77.25-90.30) | 84.80 (77.00-92.60) | 85.30 (78.00-92.00) | 86.00 (78.50-93.20) | 86.80 (82.00-90.20) | 89.00 (81.00-95.00) | <0.001 |
| LDL-C, mg/dL | | 112.50 (92.40-134.15) | 116.95 (97.81-139.95) | 109.79 (93.17-134.54) | 111.34 (88.53-137.44) | 119.65 (96.26-144.20) | 122.17 (95.10-143.43) | 106.51 (84.67-129.51) | <0.001 |
| HDL-C, mg/dL | | 50.64 (41.37-60.70) | 51.42 (41.37-62.05) | 49.87 (39.82-59.92) | 49.87 (39.82-61.08) | 48.52 (40.59-58.38) | 54.12 (42.14-63.02) | 47.17 (39.05-59.92) | <0.249 |
| CRP, mg/L | | 0.88 (0.50-1.85) | 0.94 (0.52-1.80) | 0.71 (0.47-1.08) | 1.03 (0.54-2.40) | 1.09 (0.58-2.15) | 1.07 (0.56-1.66) | 1.22 (0.61-2.49) | 0.002 |
| The TyG index | | 8.51 (8.16-8.91) | 8.56 (8.22-8.96) | 8.73 (8.36-9.14) | 8.63 (8.16-9.01) | 8.67 (8.29-9.04) | 8.55 (8.18-8.98) | 8.77 (8.40-9.31) | <0.001 |

Note: WC, waist circumstance. LDL-C, low-density lipoprotein cholesterol. HDL-C, high-density lipoprotein cholesterol. CRP, C-reactive protein. The TyG index, triglyceride-glucose index. Values are presented as number (N) with percent (%) for categorical variables or median with interquartile range (IQR) for continuous variables.

### Table S11. Association between TyG index in 2011 and progressions of hypertension stages from 2011 to 2015

|  |  | **Normotension to Stage 1** | **Normotension to Stage 2** | **Maintained Stage 1** | **Stage 1 to Stage 2** | **Maintained Stage 2** |
| --- | --- | --- | --- | --- | --- | --- |
|  |  | **Adjusted OR (95% CI)** | | | | |
| ***Overall (N=4,144)*** | | | |  |  |  |
|  | Quartile 1 | Reference | Reference | Reference | Reference | Reference |
|  | Quartile 2 | 1.10 (0.83-1.45) | 1.63 (0.91-2.93) | 1.23 (0.85-1.79) | 1.25 (0.64-2.44) | 1.22 (0.58-2.57) |
|  | Quartile 3 | 1.40 (1.05-1.86) | 0.84 (0.41-1.74) | 1.11 (0.74-1.66) | 1.33 (0.66-2.70) | 1.97 (0.96-4.03) |
|  | Quartile 4 | 1.45 (1.05-2.00) | 1.33 (0.64-2.78) | 1.68 (1.10-2.56) | 2.08 (1.00-4.35) | 2.62 (1.23-5.62) |
|  | TyG-continuous | 1.39 (1.16-1.65) | 1.05 (0.70-1.55) | 1.32 (1.05-1.67) | 1.34 (0.90-2.00) | 2.00 (1.37-2.91) |
| ***Male (N=1,946)*** | | | |  |  |  |
|  | Quartile 1 | Reference | Reference | Reference | Reference | Reference |
|  | Quartile 2 | 1.02 (0.70-1.49) | 1.05 (0.46-2.39) | 1.00 (0.58-1.73) | 1.22 (0.53-2.78) | 1.15 (0.45-2.89) |
|  | Quartile 3 | 1.50 (1.01-2.23) | 0.75 (0.28-2.02) | 1.18 (0.65-2.13) | 1.53 (0.64-3.66) | 1.48 (0.55-4.01) |
|  | Quartile 4 | 1.54 (0.99-2.40) | 0.77 (0.25-2.32) | 1.81 (0.99-3.30) | 2.00 (0.79-5.06) | 1.08 (0.34-3.41) |
|  | TyG-continuous | 1.53 (1.20-1.94) | 0.91 (0.50-1.67) | 1.34 (0.95-1.90) | 1.33 (0.81-2.21) | 1.18 (0.64-2.18) |
| ***Female (N=2,198)*** | | | |  |  |  |
|  | Quartile 1 | Reference | Reference | Reference | Reference | Reference |
|  | Quartile 2 | 1.16 (0.76-1.76) | 2.60 (1.09-6.24) | 1.41 (0.83-2.39) | 1.27 (0.40-4.00) | 1.58 (0.43-5.85) |
|  | Quartile 3 | 1.26 (0.82-1.94) | 1.06 (0.36-3.13) | 1.03 (0.58-1.82) | 0.92 (0.27-3.19) | 3.23 (1.01-10.47) |
|  | Quartile 4 | 1.23 (0.75-1.99) | 2.49 (0.86-7.21) | 1.57 (1.05-2.88) | 2.11 (0.60-7.51) | 5.03 (1.48-17.14) |
|  | TyG-continuous | 1.16 (0.89-1.51) | 1.21 (0.71-2.06) | 1.30 (0.94-1.79) | 1.39 (0.68-2.84) | 2.52 (1.51-4.19) |

Note: TyG, triglyceride-glucose. OR, odds ratio. CI, confidence interval. OR values were adjusted for age, sex, residence, education, economic status, tobacco use, alcohol consumption, general obesity, waist circumstance, low-density lipoprotein cholesterol, high-density lipoprotein cholesterol, and C-reactive protein.

### Table S12. Association between TyG index in 2011 and progressions of hypertension phenotypes from 2011 to 2015

|  |  | **Normotension to ISH** | **Normotension to IDH** | **Normotension to SDH** | **Maintained ISH** | **ISH to SDH** | **Maintained SDH** |
| --- | --- | --- | --- | --- | --- | --- | --- |
|  |  | **Adjusted OR (95% CI)** | | | | | |
| ***Overall (N=4,126)*** | | | |  |  |  |  |
|  | Quartile 1 | Reference | Reference | Reference | Reference | Reference | Reference |
|  | Quartile 2 | 1.26 (0.93-1.70) | 2.07 (0.90-4.78) | 0.74 (0.43-1.29) | 1.26 (0.84-1.89) | 0.97 (0.47-1.99) | 1.20 (0.62-2.32) |
|  | Quartile 3 | 1.26 (0.91-1.74) | 2.72 (1.15-6.40) | 1.19 (0.69-2.03) | 1.57 (1.04-2.37) | 0.84 (0.37-1.88) | 1.15 (0.56-2.37) |
|  | Quartile 4 | 1.33 (0.92-1.92) | 3.46 (1.42-8.44) | 1.24 (0.69-2.22) | 1.63 (1.03-2.57) | 1.23 (0.53-2.84) | 2.66 (1.36-5.20) |
|  | TyG-continuous | 1.28 (1.04-1.56) | 1.94 (1.27-2.97) | 1.24 (0.89-1.72) | 1.34 (1.05-1.70) | 1.20 (0.73-1.98) | 1.71 (1.22-2.40) |
| ***Male (N=1,934)*** | | | |  |  |  |  |
|  | Quartile 1 | Reference | Reference | Reference | Reference | Reference | Reference |
|  | Quartile 2 | 1.21 (0.82-1.82) | 1.74 (0.49-6.16) | 0.51 (0.24-1.10) | 1.17 (0.65-2.09) | 0.59 (0.24-1.46) | 1.09 (0.45-2.61) |
|  | Quartile 3 | 1.52 (0.97-2.38) | 2.48 (0.70-8.81) | 0.94 (0.45-1.96) | 1.80 (0.98-3.30) | 0.53 (0.18-1.51) | 0.98 (0.36-2.69) |
|  | Quartile 4 | 1.25 (0.74-2.11) | 3.70 (1.02-13.44) | 1.26 (0.60-2.64) | 1.65 (0.83-3.28) | 0.42 (0.12-1.42) | 3.24 (1.38-7.63) |
|  | TyG-continuous | 1.39 (1.04-1.84) | 2.38 (1.29-4.39) | 1.26 (0.83-1.91) | 1.32 (0.90-1.95) | 0.57 (0.28-1.17) | 1.93 (1.23-3.01) |
| ***Female (N=2,192)*** | | | |  |  |  |  |
|  | Quartile 1 | Reference | Reference | Reference | Reference | Reference | Reference |
|  | Quartile 2 | 1.27 (0.81-2.01) | 2.29 (0.74-7.10) | 1.11 (0.48-2.56) | 1.33 (0.75-2.37) | 3.48 (0.69-17.43) | 1.30 (0.46-3.63) |
|  | Quartile 3 | 1.03 (0.63-1.67) | 3.13 (0.97-10.12) | 1.43 (0.62-3.31) | 1.39 (0.78-2.48) | 3.20 (0.59-17.28) | 1.33 (0.46-3.85) |
|  | Quartile 4 | 1.23 (0.72-2.09) | 3.64 (1.01-13.08) | 0.98 (0.37-2.63) | 1.47 (0.78-2.80) | 6.89 (1.21-39.24) | 1.81 (0.59-5.56) |
|  | TyG-continuous | 1.11 (0.83-1.48) | 1.82 (0.96-3.45) | 1.02 (0.60-1.73) | 1.28 (0.92-1.78) | 2.60 (1.26-5.38) | 1.40 (0.80-2.43) |

Note: TyG, triglyceride-glucose. ISH, isolated systolic hypertension. IDH, isolated diastolic hypertension. SDH, systolic diastolic hypertension. OR, odds ratio. CI, confidence interval. OR values were adjusted for age, sex, residence, education, economic status, tobacco use, alcohol consumption, general obesity, waist circumstance, low-density lipoprotein cholesterol, high-density lipoprotein cholesterol, and C-reactive protein.
